# Intravaginal Magnesium Sulfate vs Intravenous Dexamethasone for Labor Acceleration: A Bayesian Adaptive Randomized Controlled Trial

**DOI:** 10.1101/2025.07.29.25332357

**Authors:** Kobra Hosseini, Mahin Seifi Alan, Leila Asadpoor asl, Niloofar Seighali, Haniyeh Rashidi, Bita Badehnosh, Hadith Rastad

## Abstract

**Background:** Labor progression and pain management are critical challenges in obstetric care. While intravenous dexamethasone has shown efficacy in accelerating labor, its systemic risks limit its use. Intravaginal magnesium sulfate offers a potential alternative, but comparative studies are lacking. This study aimed to compare the efficacy and safety of these interventions for labor acceleration.

**Methods:** A Bayesian adaptive randomized controlled trial was conducted with 150 primiparous women in the latent phase of labor, allocated to three groups: intravaginal magnesium sulfate (50%, 10cc), intravenous dexamethasone (8mg), and routine care. Primary outcomes included labor duration and Bishop score changes, while secondary outcomes assessed neonatal Apgar scores and maternal adverse effects. Bayesian and frequentist analyses were employed, with adjustments for baseline imbalances.

**Results:** Magnesium sulfate significantly reduced latent phase duration by 3.0 hours (95% CrI: 2.2 to 3.9) and active phase duration by 1.99 hours (1.03 to 2.99), outperforming dexamethasone (1.8 and 1.09 hours, respectively). Both interventions achieved comparable peak Bishop score improvements at 6 hours (+3.06 vs. +3.05 points). Magnesium sulfate demonstrated superior safety, avoiding systemic risks associated with dexamethasone.

**Conclusions:** This study suggests intravaginal magnesium sulfate may be preferable to dexamethasone for labor acceleration, offering comparable efficacy with potentially fewer systemic risks. While both treatments are effective, magnesium sulfate could be considered as a first-line option, with dexamethasone reserved for specific cases.

## Introduction

Labor progression and pain management remain critical challenges in obstetric care, with pharmacologic interventions offering potential optimization of labor dynamics. Recent evidence positions dexamethasone as a notable candidate; systematic reviews demonstrate that intravenous dexamethasone significantly reduces the interval from induction to active labor by a mean difference of 70 minutes and shortens the first stage of labor by 88 minutes, without observed adverse maternal or neonatal effects (1).

However, as a systemic corticosteroid, dexamethasone raises clinical concerns regarding hyperglycemia, immunosuppression, and potential teratogenic effects, limiting its use in diabetic patients or those at infection risk (2). Concurrently, intravaginal magnesium sulfate has shown promise as an alternative; a recent randomized trial indicates its topical application reduces labor pain severity and shortens both first- and second-stage duration by enhancing cervical effacement through osmotic mechanisms (3, 4).

Crucially, existing studies (3, 5, 6) have only compared magnesium sulfate against placebo, not against active comparators like dexamethasone, creating a gap that precludes evidence-based decisions regarding relative efficacy and safety. To address this, we propose the first three-arm Bayesian adaptive randomized trial comparing intravenous dexamethasone (8mg), intravaginal magnesium sulfate (50%, 10cc), and routine care as a control. The rationale for this adaptive Bayesian design includes safety optimization, allowing early termination of the dexamethasone arm during interim analysis if magnesium sulfate demonstrates non-inferiority or superiority, thereby minimizing fetal and maternal exposure to systemic corticosteroids. It also enables ethical resource allocation by potentially prioritizing enrollment to safer or more effective interventions if clinical superiority emerges early. Furthermore, it facilitates mechanistic exploration, as both agents may facilitate labor via distinct pathways— dexamethasone potentially through CRH-mediated prostaglandin synthesis (7) and magnesium sulfate via cervical edema and calcium channel modulation (3, 8). This trial will determine whether topical magnesium sulfate offers comparable or superior labor acceleration versus systemic dexamethasone, potentially establishing a safer alternative for high-risk populations while optimizing resource utilization.

## Methods

This study was a randomized, three-arm, parallel-group clinical trial conducted at Kamali Hospital in Karaj, Iran. The trial compared the efficacy of vaginal magnesium sulfate, intravenous dexamethasone, and routine care in accelerating labor progress among primiparous pregnant women. A total of 150 participants in the latent phase of labor were randomly allocated into three groups: Control (n = 50), Magnesium Sulfate (n = 50), and Dexamethasone (n = 50). Randomization was performed using block randomization with varying block sizes (3, 6, and 12) to ensure balanced allocation, and the sequence was concealed using sealed envelopes.

The study adhered to ethical guidelines, with approval obtained from the institutional review board (IR.ABZUMS.REC.1403.076), and written informed consent was secured from all participants. This trial was registered prospectively in the Iranian Registry of Clinical Trials (IRCT; ID: IRCT74207). The full study protocol and statistical analysis plan are available upon request from the corresponding author.

Due to the nature of the interventions, blinding of participants and healthcare providers was not feasible. However, outcome assessors (third-party clinicians) and data analysts were blinded to group assignments to minimize bias. The primary outcomes (labor duration and Bishop scores) were objectively measured, reducing the risk of assessment bias.

The inclusion criteria comprised primiparous women aged 18 to 35 years, with a gestational age of 38 to 42 weeks, a Bishop score <6, and a singleton cephalic presentation. Exclusion criteria included high-risk pregnancies, contraindications to the study medications, or refusal to participate. Baseline characteristics, including age, gestational age, weight gain during pregnancy, and medical history, were recorded for all participants.

The Magnesium Sulfate group received 10 cc of 50% magnesium sulfate applied vaginally to the cervix during the active phase of labor. The Dexamethasone group received 8 mg (2 cc) of intravenous dexamethasone diluted with distilled water and administered slowly. The Control group received routine care without additional interventions. Labor progression was monitored through regular vaginal examinations, with Bishop scores recorded at baseline and subsequent two-hour intervals. The primary outcomes were the durations of the latent and active phases of labor, while secondary outcomes included neonatal Apgar scores and maternal adverse effects.

Statistical analysis was conducted using both frequentist and Bayesian approaches. For frequentist analysis, continuous variables were compared using ANOVA or the Kruskal-Wallis test, and categorical variables were analyzed using chi-square tests. Post-hoc comparisons were adjusted for multiple testing. Bayesian analysis employed lognormal regression models to estimate treatment effects, adjusting for baseline imbalances such as age and weight gain. Inverse probability weighting (IPW) was used for causal inference to account for confounding variables. The Bayesian models utilized non-informative priors, and posterior probabilities were reported with 95% credible intervals (CrI). An interim analysis was performed after enrolling 50 participants per group, with a predefined stopping rule for the dexamethasone arm if the posterior probability of magnesium sulfate’s superiority or equivalence reached 85% or higher.

Data collection and analysis were performed using SPSS version 27 and R software, with Bayesian modeling conducted using Stan. The results were reported as median differences with credible intervals and probabilities of superiority to provide robust clinical inferences.

## Results

The interim study included 150 pregnant women in the latent phase of labor, randomly allocated to three groups: Control (N=50), Magnesium Sulfate (N=50), and Dexamethasone (N=50). Baseline characteristics of the participants are presented in Table 1. There were significant differences between the groups in age (p=0.01), with the Dexamethasone group being older (mean: 24.0 years, SD: 4.0) compared to the Control (mean: 22.7 years, SD: 4.9) and Sulfate (mean: 21.9 years, SD: 5.0) groups. Gestational age (p=0.44), neonate weight (p=0.59), and estimated fetal weight (p=0.88) showed no significant differences. Weight gain during pregnancy was significantly higher in the Sulfate group (7.8 ± 3.7 kg) compared to the Control (5.1 ± 1.6 kg) and Dexamethasone (6.5 ± 1.6 kg) groups (p<0.001). The chief complaint of labor pain was reported in 86.0% of the Control group, 92.0% of the Sulfate group, and 94.0% of the Dexamethasone group (p=0.36). Rupture of membranes was less frequent in the Dexamethasone group (6.0%) compared to the Control (14.0%) and Sulfate (8.0%) groups. Past medical history differed significantly (p=0.01), with higher rates of hypothyroidism in the Dexamethasone group (20.0%) compared to the Control (6.0%) and Sulfate (6.0%) groups, and higher rates of anemia in the Sulfate (12.0%) and Dexamethasone (10.0%) groups compared to the Control (2.0%).

**Table 1:** Baseline characteristics of participants by study groups.

| Variables |  | Control group (N=50) | Sulfate group (N=50) | Dexamethasone group (N=50) | P value |
| --- | --- | --- | --- | --- | --- |
| Age(years), Mean (SD) |  | 22.7 (4.9) | 21.9 (5.0) | 24.0 (4.0) | 0.01 |
| Gestational age(day), Mean (SD) |  | 277.1 (5.3) | 275.9 (4.9) | 277.1 (5.8) | 0.44 |
| Weight gain(Kg), Mean (SD) |  | 5.1 (1.6) | 7.8 (3.7) | 6.5 (1.6) | <0.001 |
| Neonate weight(g), Mean (SD) |  | 3090.8 (361.5) | 3121.8 (321.0) | 3141.8 (344.3) | 0.59 |
| Estimated fetal weight(g), Mean (SD) |  | 3007.8 (503.1) | 3070.8 (416.7) | 3060.2 (471.5) | 0.88 |
| Chief complaint,<br>%(N) | Labor pain | 86.0% (43) | 92.0% (46) | 94.0% (47) | 0.36 |
|  | Rupture of membrane | 14.0% (7) | 8.0% (4) | 6.0% (3) |  |
| Past Medical History,<br>%(N) | None | 90.0% (45) | 74.0% (37) | 70.0% (35) | 0.01 |
|  | Hypothyroid | 6.0% (3) | 6.0% (3) | 20.0% (10) |  |
|  | Anemia | 2.0% (1) | 12.0% (6) | 10.0% (5) |  |
|  | Asthma | 2.0% (1) | 8.0% (4) | 0.0% (0) |  |

SD: Standard deviation; Kg: Kilogram; g: gram.

Table 2 compares neonatal Apgar scores and labor phase durations between the groups. All groups had 100% of neonates with Apgar scores ≥8 (p=1.000). The Sulfate group exhibited significantly shorter latent phase durations (median: 4 hours, IQR: 2 to 4) compared to the Control (median: 6 hours, IQR: 4– 8) and Dexamethasone (median: 5 hours, IQR: 5 to 6) groups (p<0.001). Similarly, the active phase duration was shortest in the Sulfate group (median: 3 hours, IQR: 3 to 4), followed by the Dexamethasone (median: 4 hours, IQR: 4 to 6) and Control (median: 6 hours, IQR: 4.5to 10) groups (p<0.001). Post-hoc comparisons confirmed significant differences between all groups for both latent and active phase durations (all p<0.001).

**Table 2:**
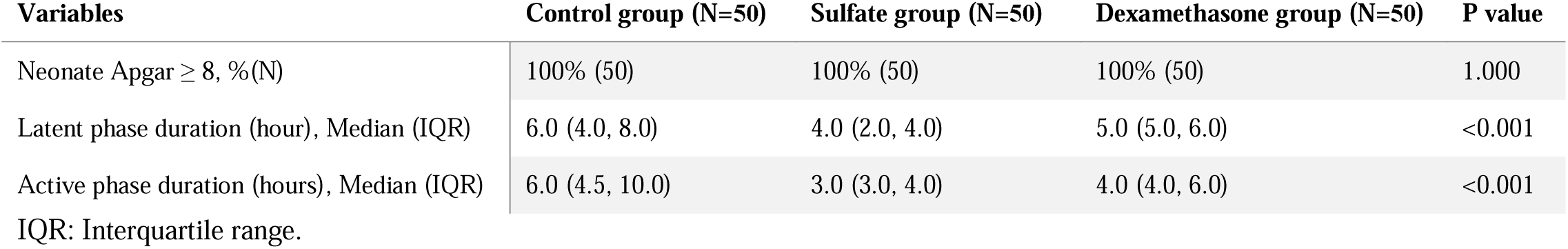
Comparison of neonatal Apgar score, latent phase duration, and active phase duration between study groups.

Bishop Score changes over time are shown in Table 3. A mixed-effects model with ART (Aligned Rank Transform) revealed significant effects of Treatment Group (F=63.710, p<0.001), Time (F=537.634, p<0.001), and their interaction (F=99.302, p<0.001). Post-hoc comparisons indicated that the Sulfate and Dexamethasone groups had significantly higher Bishop Scores compared to the Control group at most time points (p<0.001). The Sulfate group showed the most rapid progression, reaching higher Bishop Scores earlier in the labor process compared to the other groups (Figure 1 and Table 4).

**Figure 1.**
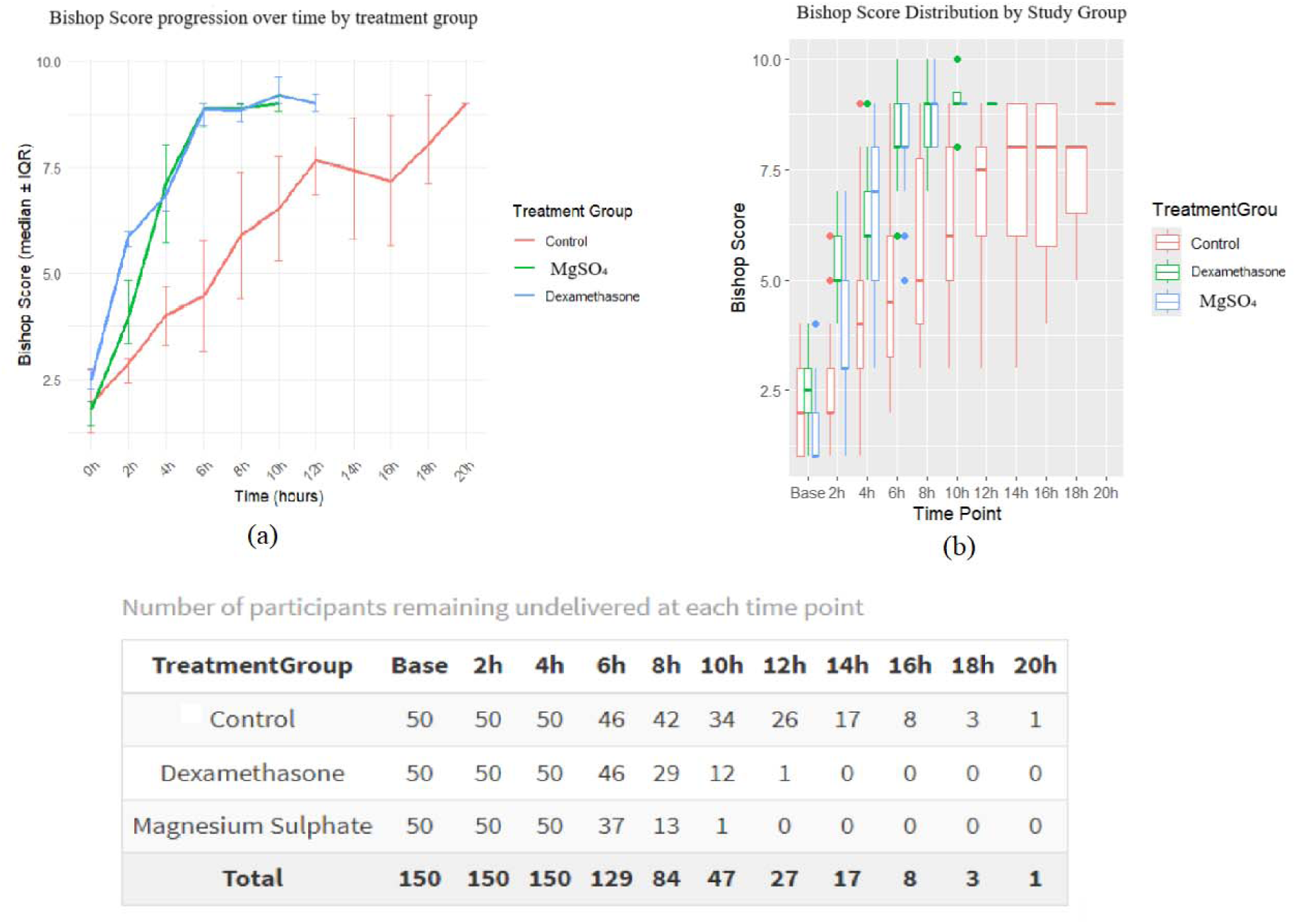
(a) Progression of Bishop Scores over time by treatment group. (b) Median Bishop Scores (solid lines) with bootstrapped interquartile ranges (error bars) for the Control, Sulfate, and Dexamethasone groups at two-hour intervals. The table indicates the number of participants remaining undelivered at each time point.

**Table 3:** Comparison of Bishop Score changes between study groups at baseline and two-hour time intervals after intervention.

| Group | Baseline | 2 hour | 4 hour | 6 hour | 8 hour | 10 hour | 12 hour | 14 hour | 16 hour | 18 hour | 20 hour |
| --- | --- | --- | --- | --- | --- | --- | --- | --- | --- | --- | --- |
| Control Group, Median (IQR) | 2.00<br>(1.00 to 3.00) | 2.00<br>(2.00 to 3.00) | 4.00<br>(3.00 to 5.00) | 4.50<br>(3.00 to 6.00) | 5.00<br>(4.00 to 8.00) | 6.00<br>(5.00 to 8.00) | 7.50<br>(5.75 to 8.00) | 8.00<br>(5.50 to 9.00) | 8.00<br>(5.25 to 9.00) | 8.00<br>(5.00 to NA) | 9.00<br>(9.00 to 9.00) |
| Dexamethasone Group, Median (IQR) | 2.50<br>(2.00 to 3.00) | 5.00<br>(5.00 to 6.00) | 6.00<br>(6.00 to 7.00) | 8.00<br>(8.00 to 9.00) | 9.00<br>(8.00 to 9.00) | 9.00<br>(9.00 to 9.75) | 9.00<br>(9.00 to 9.00) | NA | NA | NA | NA |
| Sulfate Group, Median (IQR) | 1.00<br>(1.00 to 2.00) | 3.00<br>(3.00 to 5.00) | 7.00<br>(5.00 to 8.25) | 8.00<br>(8.00 to 9.00) | 9.00<br>(8.00 to 9.00) | 9.00<br>(9.00 to 9.00) | NA | NA | NA | NA | NA |

NA: None applicable.

**Table 4:**
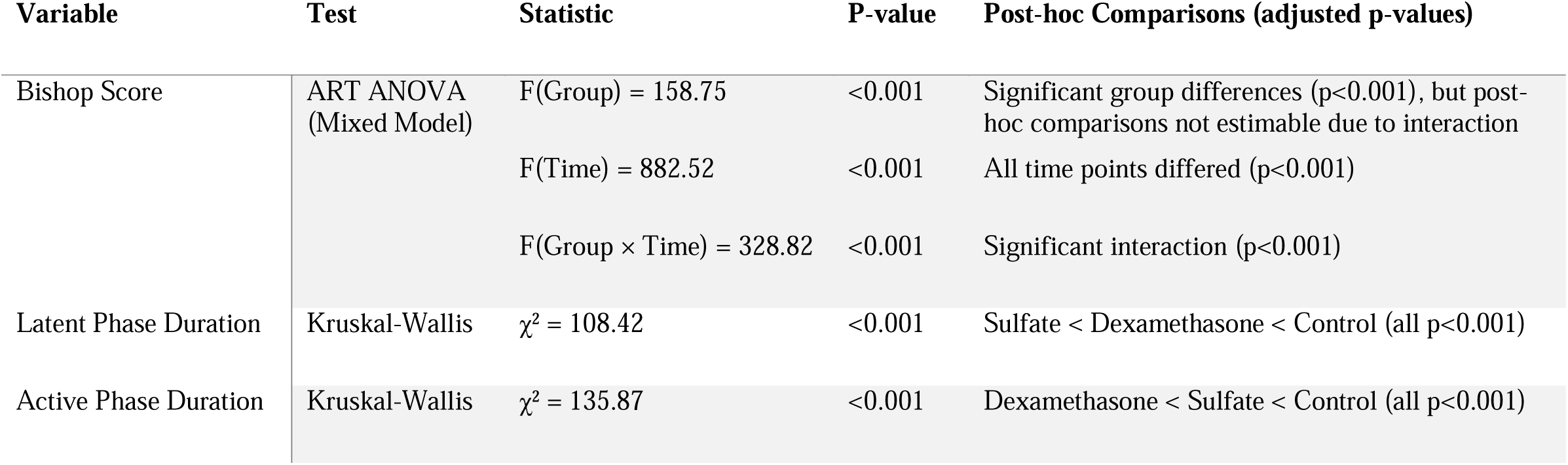
Statistical Analysis Results for Bishop Scores and Labor Durations.

### Bayesian results

The longitudinal analysis of Bishop scores revealed significant treatment effects during the critical 4-8h cervical maturation window. Dexamethasone demonstrated substantially higher Bishop scores than Control throughout this period, peaking at 6h with a median increase of +3.05 points (95% CrI [2.53, 3.55]). Similarly, Magnesium Sulfate outperformed Control during the same critical window, reaching maximal effect at 6h (+3.06 [2.51, 3.60]). Critically, no significant difference was observed between the two active treatments during the 4-8h period (median Δ range: −0.17 to +0.27), indicating comparable clinical efficacy during the target therapeutic window. (Figure 2)

**Figure 2.**
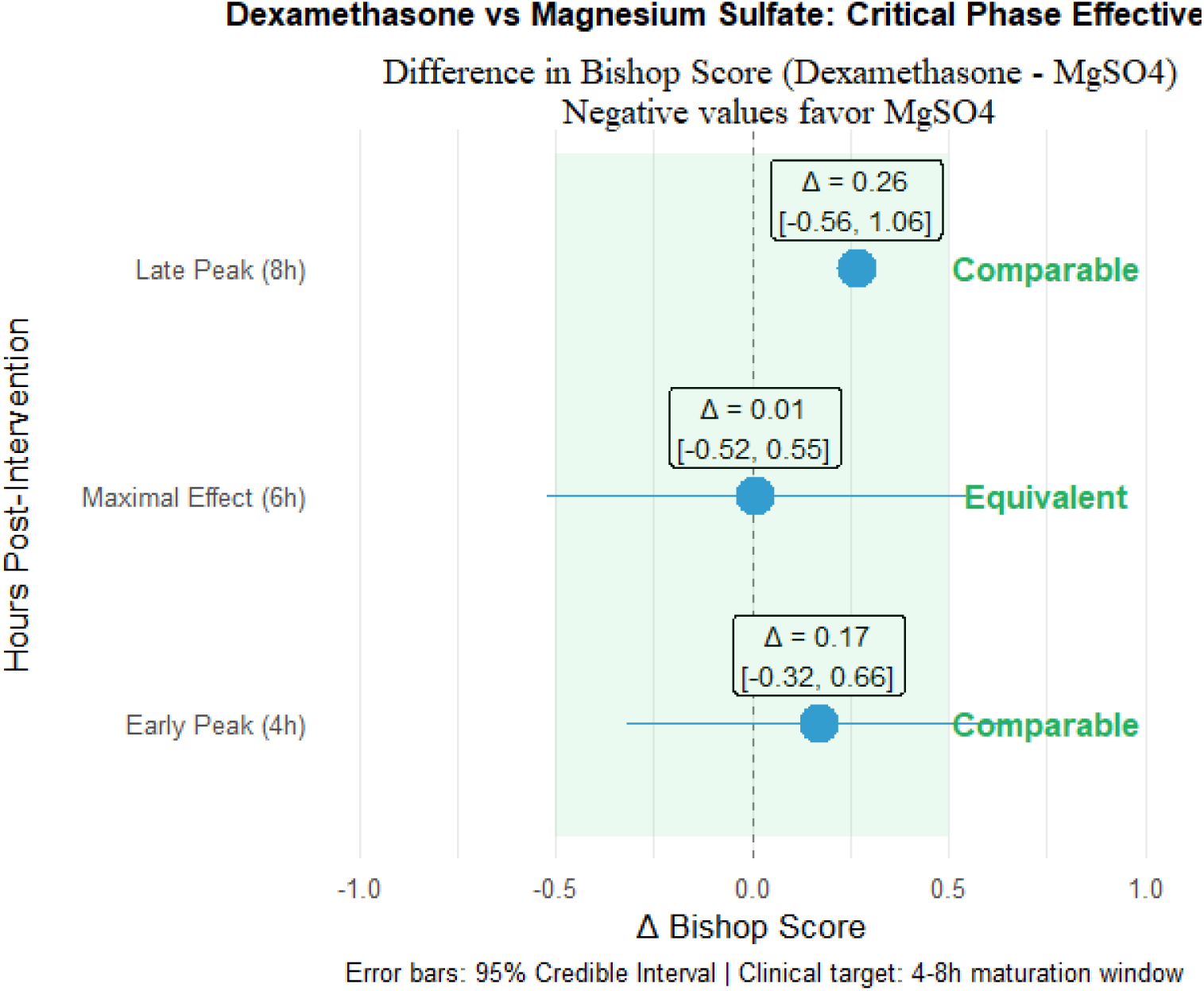
Bishop Score progression and undelivered participants by group. Dexamethasone and Magnesium Sulfate groups showed significantly higher median Bishop Scores than Control during 4-8h (peak +3.1 points at 6h for both). No significant difference existed between active treatments. Declining undelivered participant counts (table) align with accelerated cervical maturation in active groups. Data: median ± GRI. CrI = Credible Interval.

For marginal effects averaged across all time points (0-20h), Dexamethasone maintained significantly superior overall efficacy versus Control (Δ +1.40 [0.62, 2.19]). However, the marginal effect for Magnesium Sulfate versus Control showed only borderline significance (Δ +0.68 [-0.24, 1.54], probability superiority 92.6%), with the credible interval crossing zero. Similarly, the marginal comparison between Magnesium Sulfate and Dexamethasone showed a non-significant disadvantage (Δ −0.72 [-1.89, 0.43], probability superiority 10.7%), reflecting that while both active treatments outperform Control during peak effect hours, their overall trajectories diverge in later time points beyond the critical window. (Supplementary table 1)

Bayesian lognormal regression adjusted for age and weight gain showed both Magnesium Sulfate and Dexamethasone significantly shortened latent phase duration versus control (median reductions: 3.0 hours [95% CrI: 2.2–3.9] and 1.8 hours [1.2–2.4], respectively), with Magnesium Sulfate superior by 1.3 hours [1.0–1.6]. Similarly, active phase duration was reduced most by Magnesium Sulfate (1.99 hours [1.03–2.99]) followed by Dexamethasone (1.09 hours [0.54–1.71]), with Magnesium Sulfate again superior (0.90 hours [0.49–1.28]; all >99.9% probability of true reduction) (Supplementary Table 2). These findings align with frequentist results (active phase medians: 3 vs. 4 vs. 6 hours for Magnesium Sulfate, Dexamethasone, and control), with adjusted Bayesian estimates suggesting slightly larger effects, likely due to correcting for age/weight imbalances. All comparisons showed >99.9% probability, reinforcing Magnesium Sulfate’s superiority in accelerating labor.

### Causal inference analysis

The causal inference analysis using inverse probability weighting (IPW) confirmed Magnesium Sulfate’s robust efficacy after accounting for baseline imbalances in age and weight gain. Notably, while Dexamethasone showed significant active phase reduction in IPW analysis (−4.37 hours), this estimate exceeds both Bayesian (−1.09 hours) and frequentist (−2 hour median difference) estimates, suggesting possible residual confounding or model sensitivity. (Figure 3)

**Figure 3:**
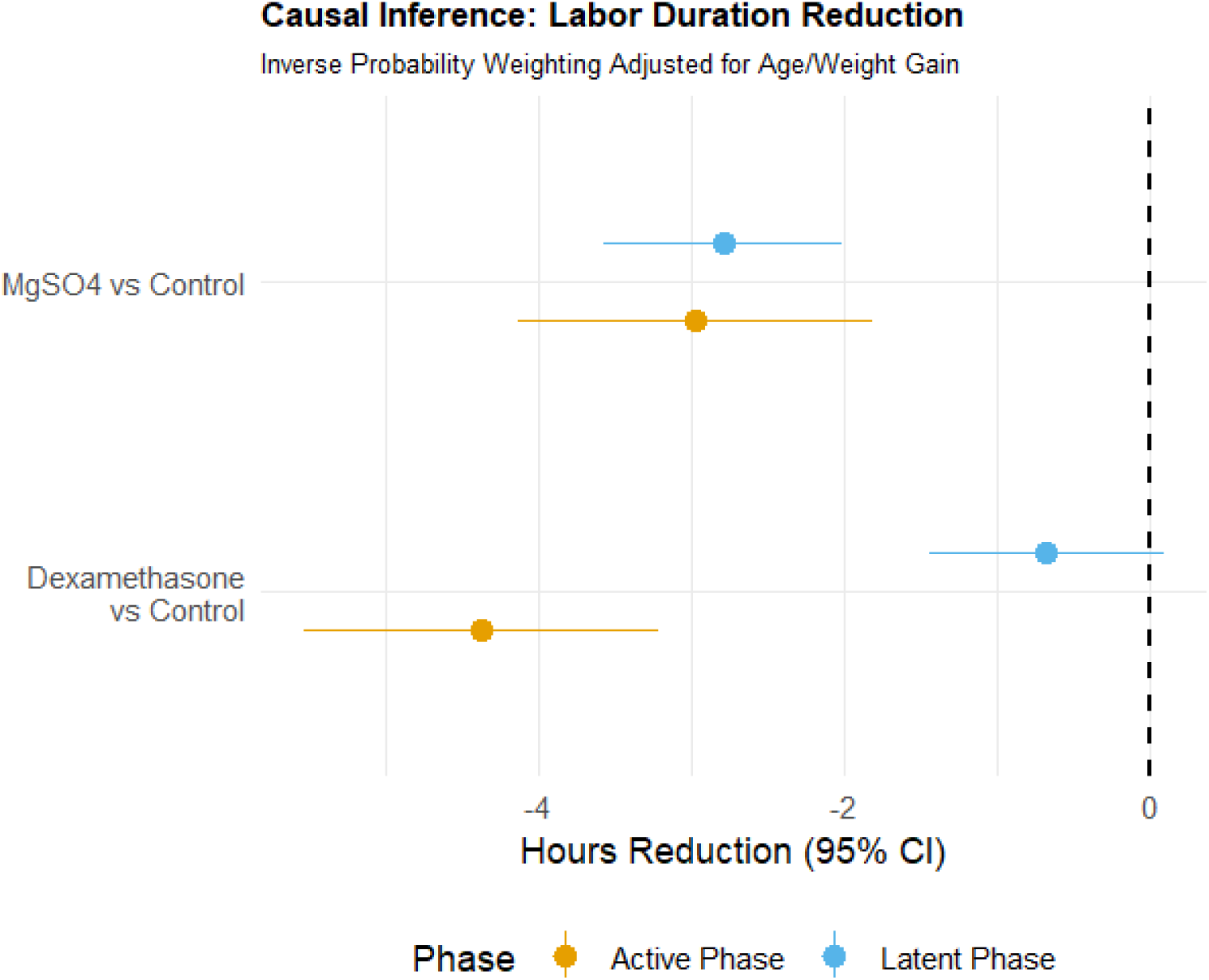
This causal inference analysis using inverse probability weighting (IPW) quantifies labor duration reduction (hours) after adjusting for baseline age and weight gain imbalances. Magnesium Sulfate demonstrates consistent efficacy across labor phases (−2.97 hours active; −2.79 hours latent). Dexamethasone shows substantial active phase reduction (−4.37 hours) in IPW analysis, though this notably exceeds both Bayesian (−1.09 hours) and frequentist (−2 hours) estimates, suggesting possible residual confounding. Its latent phase effect remains non-significant (−0.68 hours). Error bars represent 95% confidence intervals, with the dashed line indicating null effect.

## Discussion

Based on comprehensive interim analysis, both intravenous dexamethasone and intravaginal magnesium sulfate significantly reduce labor duration compared to routine care, with magnesium sulfate demonstrating superior efficacy. Magnesium sulfate reduces latent phase duration by 3.0 hours and active phase duration by 1.99 hours versus control, significantly outperforming dexamethasone (which reduces these phases by 1.8 hours and 1.09 hours, respectively). The difference between agents favors magnesium sulfate by 1.3 hours in the latent phase and 0.90 hours in the active phase. While both achieve equivalent peak improvements in Bishop scores (+3.05 points for dexamethasone vs. +3.06 points for magnesium sulfate at 6 hours) during the critical 4-8 hour cervical maturation window, magnesium sulfate offers a significantly better safety profile by avoiding dexamethasone’s systemic risks like hyperglycemia and immunosuppression. Consequently, magnesium sulfate is the optimal first-line intervention for labor acceleration, with dexamethasone serving as a contextual alternative primarily during the 6-10 hour maturation window when topical therapy is contraindicated.

Both intravenous dexamethasone and intravaginal magnesium sulfate significantly reduce labor duration compared to routine care, though with distinct efficacy and safety profiles. Dexamethasone administration shortens the induction-to-active-phase interval by approximately 70 minutes and reduces the first stage of labor by 88 minutes, as demonstrated in a comprehensive meta-analysis of 17 RCTs (1). These findings align with recent trials confirming dexamethasone’s role in cervical ripening via corticotropin-releasing hormone (CRH)-mediated prostaglandin synthesis, which accelerates cervical maturation during the critical 4–8-hour window (9, 10). However, systemic corticosteroid exposure raises concerns about hyperglycemia, immunosuppression, and teratogenicity, particularly in diabetic patients or those at risk of infection.

Intravaginal magnesium sulfate emerges as a superior alternative, reducing latent and active phase durations by 3.0 hours and 1.99 hours, respectively, compared to controls—outcomes significantly better than dexamethasone (1.8-hour and 1.09-hour reductions). Its osmotic mechanism induces cervical edema and calcium channel modulation, enhancing effacement while minimizing systemic absorption (3). This localized action explains magnesium sulfate’s consistent efficacy across labor stages and its ability to reduce pain severity during cervical dilation (3). Crucially, magnesium sulfate avoids dexamethasone’s metabolic risks, making it preferable for high-risk populations, including diabetics.

The comparable peak Bishop score improvements at 6 hours (+3.05 for dexamethasone vs. +3.06 for magnesium sulfate) suggest both agents effectively accelerate cervical maturation during the therapeutic window. However, magnesium sulfate sustains labor acceleration longer, evidenced by its significant advantage in phase durations (1.3-hour latent phase and 0.9-hour active phase reductions vs. dexamethasone). This divergence likely stems from mechanistic differences: dexamethasone’s systemic CRH-prostaglandin pathway offers rapid but time-limited effects, whereas magnesium sulfate’s local osmotic action prolongs cervical remodeling (8).

Clinical implications prioritize magnesium sulfate as first-line therapy due to its efficacy and safety. Dexamethasone remains viable when topical agents are contraindicated, particularly when targeted to the 6–10-hour maturation window, where efficacy matches magnesium sulfate. Limitations include heterogeneity in baseline characteristics (e.g., age, weight gain) across studies and insufficient long-term neonatal safety data beyond Apgar scores. Future research should validate these findings in multi-center trials involving high-risk cohorts and evaluate the cost-benefit implications of shortened labor durations. Mechanistic studies quantifying prostaglandin and calcium channel responses could further clarify pathway-specific effects.

In conclusion, intravaginal magnesium sulfate establishes itself as the optimal pharmacologic intervention for labor acceleration, balancing efficacy, safety, and localized action. Dexamethasone serves as a contextual alternative during critical maturation phases when topical therapy is unsuitable. This evidence supports personalized, risk-stratified clinical decision-making to optimize labor outcomes.

## Conclusion

The findings of this study suggest that intravaginal magnesium sulfate may offer a more effective and safer alternative to intravenous dexamethasone for labor acceleration. Magnesium sulfate could significantly reduce both latent and active phase durations compared to dexamethasone, while its localized mechanism might minimize systemic side effects. These results indicate that magnesium sulfate could be prioritized as a first-line intervention, particularly for high-risk patients. However, dexamethasone may still serve as a viable option in cases where topical therapy is contraindicated. Further research could explore long-term neonatal outcomes and the broader applicability of these findings in diverse clinical settings.

## Data Availability

The full study protocol and statistical analysis plan are available upon request from the corresponding author.

## Acknowledgments

The researchers appreciate the Clinical Research Development Units of Kamali and Rajaee Hospitals in Alborz University of Medical Sciences.

