## Supplementary Table 1, Supplementary Table 2 for "Intravaginal Magnesium Sulfate vs Intravenous Dexamethasone for Labor Acceleration: A Bayesian Adaptive Randomized Controlled Trial"

**Supplementary Table 1:** Comparisons of Bishop Score between the three treatment groups from the Bayesian growth model

| **Pairwise comparisons** | | | | | |
| --- | --- | --- | --- | --- | --- |
| **Time Point** | **Contrast** | **value** | **lower** | **upper** | **prob_positive** |
| Base | Dexamethasone - Control | 0.102 | -0.360 | 0.577 | 0.666 |
| Base | Magnesium Sulphate - Control | -0.544 | -1.00 | -0.0895 | 0.0095 |
| Base | Magnesium Sulphate - Dexamethasone | -0.645 | -1.13 | -0.179 | 0.00375 |
| 2h | Dexamethasone - Control | 2.36 | 1.86 | 2.84 | 1 |
| 2h | Magnesium Sulphate - Control | 0.939 | 0.447 | 1.42 | 1 |
| 2h | Magnesium Sulphate - Dexamethasone | -1.42 | -1.89 | -0.937 | 0 |
| 4h | Dexamethasone - Control | 2.46 | 1.98 | 2.95 | 1 |
| 4h | Magnesium Sulphate - Control | 2.62 | 2.12 | 3.11 | 1 |
| 4h | Magnesium Sulphate - Dexamethasone | 0.171 | -0.313 | 0.642 | 0.745 |
| 6h | Dexamethasone - Control | 3.05 | 2.53 | 3.55 | 1 |
| 6h | Magnesium Sulphate - Control | 3.06 | 2.51 | 3.60 | 1 |
| 6h | Magnesium Sulphate - Dexamethasone | 0.0125 | -0.511 | 0.555 | 0.518 |
| 8h | Dexamethasone - Control | 2.81 | 2.24 | 3.42 | 1 |
| 8h | Magnesium Sulphate - Control | 3.08 | 2.29 | 3.85 | 1 |
| 8h | Magnesium Sulphate - Dexamethasone | 0.268 | -0.576 | 1.08 | 0.735 |
| 10h | Dexamethasone - Control | 2.63 | 1.82 | 3.42 | 1 |
| 10h | Magnesium Sulphate - Control | 0.971 | -1.11 | 3.07 | 0.829 |
| 10h | Magnesium Sulphate - Dexamethasone | -1.66 | -3.80 | 0.482 | 0.0745 |
| 12h | Dexamethasone - Control | 1.47 | -0.575 | 3.55 | 0.919 |
| 12h | Magnesium Sulphate - Control | -0.565 | -4.49 | 3.35 | 0.386 |
| 12h | Magnesium Sulphate - Dexamethasone | -2.08 | -6.54 | 2.38 | 0.186 |
| 14h | Dexamethasone - Control | 0.167 | -3.83 | 4.02 | 0.527 |
| 14h | Magnesium Sulphate - Control | -0.502 | -4.54 | 3.45 | 0.405 |
| 14h | Magnesium Sulphate - Dexamethasone | -0.648 | -6.31 | 4.73 | 0.412 |
| 16h | Dexamethasone - Control | 0.0852 | -3.88 | 4.04 | 0.520 |
| 16h | Magnesium Sulphate - Control | -0.593 | -4.55 | 3.40 | 0.390 |
| 16h | Magnesium Sulphate - Dexamethasone | -0.651 | -6.09 | 5.10 | 0.404 |
| 18h | Dexamethasone - Control | 0.0849 | -3.86 | 4.18 | 0.517 |
| 18h | Magnesium Sulphate - Control | -0.565 | -4.42 | 3.30 | 0.393 |
| 18h | Magnesium Sulphate - Dexamethasone | -0.695 | -6.18 | 4.65 | 0.406 |
| 20h | Dexamethasone - Control | 0.103 | -3.83 | 4.00 | 0.517 |
| 20h | Magnesium Sulphate - Control | -0.561 | -4.37 | 3.41 | 0.394 |
| **Marginal pairwise comparisons** | | | | | |
| averaged over time | **Contrast** | **value** | **lower** | **upper** | **prob_positive** |
|  | Dexamethasone - Control | 1.40 | 0.618 | 2.19 | 1 |
|  | Magnesium Sulphate - Control | 0.683 | -0.238 | 1.54 | 0.926 |
|  | Magnesium Sulphate - Dexamethasone | -0.718 | -1.89 | 0.431 | 0.107 |

**Supplementary Table 2:** Comparison of Latent and Active Phase Duration Across Study Groups

| **Comparison** | **Median Difference (hours)** | **95% CrI** | **Probability Superiority/Reduction** |
| --- | --- | --- | --- |
| **Latent Phase Duration** |  |  |  |
| Dexamethasone vs. Control | -1.75 | (-2.36, -1.22) | 100% |
| Magnesium Sulphate vs. Control | -3.03 | (-3.94, -2.20) | 100% |
| Magnesium vs. Dexamethasone | -1.28 | (-1.59, -0.97) | 100% |
| **Active Phase Duration** |  |  |  |
| Dexamethasone vs. Control | 1.09 | (0.54, 1.71) | >99.9% |
| Magnesium Sulphate vs. Control | 1.99 | (1.03, 2.99) | >99.9% |
| Magnesium Sulphate vs. Dexamethasone | 0.90 | (0.49, 1.28) | >99.9% |
